# Performance Comparison of a Flow Cytometry-based and Two Commercial Chemiluminescent Immunoassays for Detection and Quantification of Antibodies Binding to SARS-CoV-2 Spike Protein

**DOI:** 10.1101/2021.04.06.21254995

**Authors:** Arantxa Valdivia, Fabián Tarín, María Jesús Alcaraz, Paula Piñero, Ignacio Torres, Francisco Marco, Eliseo Albert, David Navarro

## Abstract

The performance of a laboratory-developed quantitative IgG/IgA flow cytometry-based immunoassay (FCI) using Jurkat T cells stably expressing full-length native S protein was compared against Elecsys® electrochemiluminiscent (ECLIA) Anti-SARS-CoV-2 S (Roche Diagnostics, Pleasanton, CA, USA), and LIAISON^®^ SARS-CoV-2 TrimericS IgG chemiluminiscent assay (CLIA) (Diasorin S.p.a, Saluggia, IT) for detection and quantitation of SARS-CoV-2-specific antibodies. A total of 225 serum/plasma specimens from 120 acute or convalescent COVID-19 individuals were included. Overall, IgG/IgA-FCI yielded the highest number of positives (n=179), followed by IgA-FCI (n=177), Roche ECLIA (n=175), IgG-FCI (n=172) and Diasorin CLIA (n=154). Positive percent agreement between FCI and compared immunoassays was highest for Roche ECLIA, ranging from 96.1% (IgG/IgA-FCI) to 97.7% (IgG-FCI), whereas negative percent agreement was higher between FCI and Diasosin CLIA, regardless of antibody isotype. A strong correlation (Rho:0.6-0.8) was found between IgG-FCI or IgA-FCI levels and antibodies quantified by Roche ECLIA and Diasorin CLIA. The trajectory of antibody levels delineated by the different immunoassays in 22 of patients with sequential specimens (≥3) was frequently discordant, with the exception of IgG and IgA determined by FCI assay and to a lesser extent antibodies quantified by Roche ECLIA and Diasorin CLIA. The data suggest that FCI may outperform Roche ECLIA and Diasorin CLIA in terms of clinical sensitivity for serological diagnosis of SARS-CoV-2 infection.

## INTRODUCTION

SARS-CoV-2 serological assays enable us to identify individuals infected either recently or in the past, assess humoral immune responses elicited by SARS-CoV-2 vaccines and infer level of susceptibility to reinfection or primary infection in vaccinees (1,2). Among SARS-CoV-2 structural components, Spike protein (S) elicits the most potent neutralizing antibodies, which are crucially involved in protecting against SARS-CoV-2 infection (3,4). The S protein, which is assembled into trimers in the viral membrane, interacts with angiotensin converting enzyme type 2 receptor (ACE2) through the receptor-binding domain (RBD) (5). Binding of RBD to ACE2 promotes cleavage of S into S1 and S2, and exposure of the fusion peptide located within S2, which eventually leads to cellular and viral membrane fusion (5). A large number of immunoassays using recombinant RBD, S1 or S2 subunits or full-length monomeric S protein as the binding antigen and returning either qualitative or semiquantitative results have been developed, evaluated in different studies and found to exhibit variable sensitivity and specificity (see 1,2,6,7 for review). A new generation of recently launched commercially-available SARS-CoV-2 immunoassays detect antibodies binding to the SARS-CoV-2 S protein in its native (trimeric) conformation or RBD and offer quantitative estimates of antibody levels (8-11), and preliminary results show increased sensitivity for detection of SARS-CoV-2 antibodies, as well as reliable estimates of serum neutralizing activity against SARS-CoV-2. In this context, a quantitative flow cytometry-based immunoassay (FCI) has been developed employing Jurkat T cells stably expressing the full-length native S protein, which is reported to be highly specific and display greater sensitivity than various comparative immunoassays targeting recombinant RBD or S subunit proteins (12). Here, we evaluated the performance of this FCI against two new-generation immunoassays: Elecsys® Anti-SARS-CoV-2 S (Roche Diagnostics, Pleasanton, CA, USA), and LIAISON^®^ SARS-CoV-2 TrimericS IgG assay (Diasorin S.p.a, Saluggia, IT), using sera from in- or outpatients with SARS-CoV-2 infection documented by RT-PCR.

## PATIENTS AND METHODS

### Patients and specimens

The current retrospective study was carried out using cryopreserved (−20°C) serum or plasma samples collected from the following four groups: (I) Convalescent COVID-19 patients, as clinically (13) and microbiologically documented by RT-PCR (14), who had been admitted to different hospital wards and eventually released. A total of 60 specimens from 35 patients, drawn at a median of 60 days (range, 8–141 days) since symptoms onset were included; (II) Acute COVID-19 patients admitted to the intensive care unit (ICU). A total of 115 specimens from 40 patients, collected at a median of 16 days (range, 2–43 days) after onset of symptoms were included; (III) Acute or convalescent COVID-19 subjects (n=45) who tested negative by rapid lateral flow immunoassay–LFIC– (INNOVITA 2019□nCoV Ab Test; Beijing Innovita Biological Technology, China), or CLIA (LIAISON^®^ SARS-CoV-2 S1/S2 IgG CLIA; DiaSorin, Saluggia, Italy, the MAGLUMI 2019-nCoV IgG SNIBE – Shenzhen New Industries Biomedical Engineering Co., Ltd, Shenzhen, China, or both) in use in our laboratory at the time of sample collection and routine testing. A total 50 specimens from this group, collected at a median of 44 days after onset of symptoms (range, 11–91) were included, of which 13 specimens tested negative by LFIC, 34 returned negative results by LIAISON assay and 7 by MAGLUMI assay; and (IV) Pre-pandemic sera obtained from unique blood donors (n=100). Specimens belonging to different groups of SARS-CoV-2-infected individuals were combined or treated individually, as appropriate for study purposes. The study was approved by the Ethics Committee of Hospital Clínico Universitario INCLIVA.

### Flow cytometry native SARS-CoV-2 S assay

FCI was carried out at the Hematology Department of Hospital General Universitario, Alicante, Spain, as previously described in detail (12). Briefly, transfected human Jurkat T-cell line (clone E6-1) stably expressing both the full-length native SARS-CoV-2 S protein and a truncated version of the human Epidermal growth factor receptor (huEGFRt) were used as the binding antigens (S-Jurkat). Non-transfected Jurkat cells (0-Jutkat) were used as controls. For each individual assay, a mixture of 50,000 0-Jurkat and 150,000 S-Jurkat cells was made in a single tube. Sera were diluted 1:50 in phosphate-buffered saline (PBS) containing 1% bovine serum albumin (BSA) and 0.02% sodium azide and incubated with the cell mixture for 30 min on ice. The cells were then spun down, washed with PBS-BSA and stained with mouse anti-human IgG-PerCP Jackson ImmunoResearch, Cambridgeshire, UK), anti-human IgA-Alexa Fluor 647 (Jackson ImmunoResearch) and anti-human EGFR (BV421) (Biolegend, San Diego, CA, USA). Samples were then washed and acquired on an Omnicyt flow cytometer (Cytognos S.L, Salamanca, Spain). A minimum of 50,000 viable events, discarding doublets and debris, were considered for the analyses. IgG or IgA antibodies bound to S proteins were identified by comparing the median fluorescence intensity (MFI) of the S-Jurkat and the 0-Jurkat cells in each sample. We established the difference between S-Jurkat and 0-Jurkat cells using the normalized MFI-ratio between EGFR and both antibody isotypes (IgG MFI-ratio and IgA MFI-ratio respectively). Data were analyzed using Infinicyt 2.0 software (Cytognos S.L.). Samples were considered positive for IgG or IgA when the normalized difference was ≥ 1.

### Commercially-available chemiluminescent SARS-CoV-2 S assays

Roche Elecsys® Anti-SARS-CoV-2 S (Roche Diagnostics, Pleasanton, CA, USA), an electrochemiluminescence sandwich immunoassay (ECLIA) that quantifies total (IgG and IgM) antibodies directed against RBD, was run on cobas® e601 modular analyzer (Roche Diagnostics, Rotkreuz, Switzerland). The assay is calibrated with the first WHO International Standard and Reference Panel for anti-SARS-CoV-2 antibody (15). The limit of detection of the assay is 0.4 U/ml and its quantification range is between 0.8 and 250.0 U/mL. Specimens yielding values >250 U/ml were diluted 1/10 and re-assayed. LIAISON® SARS-CoV-2 TrimericS IgG assay (Diasorin S.p.a, Saluggia, Italy), run on a DiaSorin LIAISON platform (DiaSorin, Stillwater, USA), measured IgG antibodies against a trimeric S-protein antigen. Samples yielding <13 AU/ml were considered negative. According to the manufacturer, the upper quantification limit of the assay is 800 AU/mL. Both assays were performed at the Microbiology Service at the Hospital Clínico Universitario, Valencia, Spain, following the instructions of the respective manufacturers.

### Statistical methods

Positive and negative percent agreement (PPA and NPA, respectively) between immunoassays were calculated using a diagnostic 2×2 test. Cohen’s Kappa statistics was used to assess the degree of concordance between qualitative results provided by the immunoassays and interpreted as previously recommended (16). The Spearman’s rank test was used to assess the correlation between continuous variables using the entire dataset (i.e. individuals with single and repeated measurements). Two-sided exact *P* values were reported. A *P* value <0.05 was considered statistically significant. The analyses were performed using SPSS version 20.0 (SPSS, Chicago, IL, USA).

## RESULTS

### Specificity of the Flow cytometry-based SARS-CoV-2 S immunoassay

In the current study, FCI was multiplexed for quantitative detection of SARS-CoV-2 IgG and IgA isotypes. Previously published data (12) found detection and quantitation of SARS-CoV-2-S-binding IgM to be less consistent and reliable. To evaluate the specificity of FCI, a total of 100 pre-pandemic sera collected from blood donors were assayed. None of these sera gave a positive signal on the FCI assay.

### Overall agreement between qualitative results provided by the immunoassays

When combining specimens from all three groups of SARS-CoV-2-infected patients, we found that IgG/IgA-FCI yielded the highest number of positives (n=179), closely followed by IgA-FCI (n=177), Roche ECLIA (n=175), and IgG-FCI (n=172) (Table 1). Diasorin CLIA returned a substantially lower number of positive results (n=154) than the former platforms. A subanalysis was next conducted including only sera (n=50) that scored negative by LFIC or CLIA assays routinely used at our laboratory at the time of testing request. As shown in Table 2, FCI (either IgG, IgA or IgG/IgA) yielded a greater number of positive results than Roche ECLIA or Diasorin CLIA.

Overall, PPA between FCI and the immunoassay compared was highest for Roche ECLIA, ranging from 96.1% (IgG/IgA-FCI) to 97.7% (IgG-FCI) (Table 3), whereas NPA was overall greater between FCI and Diasosin CLIA, regardless of the antibody isotype detected (91.4% to 97.2%). Inter-rater agreement between FCI (either IgG, IgA or IgG/IgA) and Roche ECLIA was strong (*k*=>0.8), while it was only moderate with Diasorin CLIA (*k*=>0.6-<0.8). Inter-rater agreement between results returned by Roche ECLIA and Diasorin CLIA was also moderate (*k*=0.76).

### Correlation between antibody levels measured by the immunoassays under comparison

A strong correlation (Rho ranging between 0.6 and 0.8) was found between IgG-FCI, IgA-FCI levels and total antibodies quantified by Roche ECLIA (Figure 1A and 1B, respectively) and IgG measured by Diasorin CLIA (Figure 1C and 1D, respectively).

**Figure 1.**
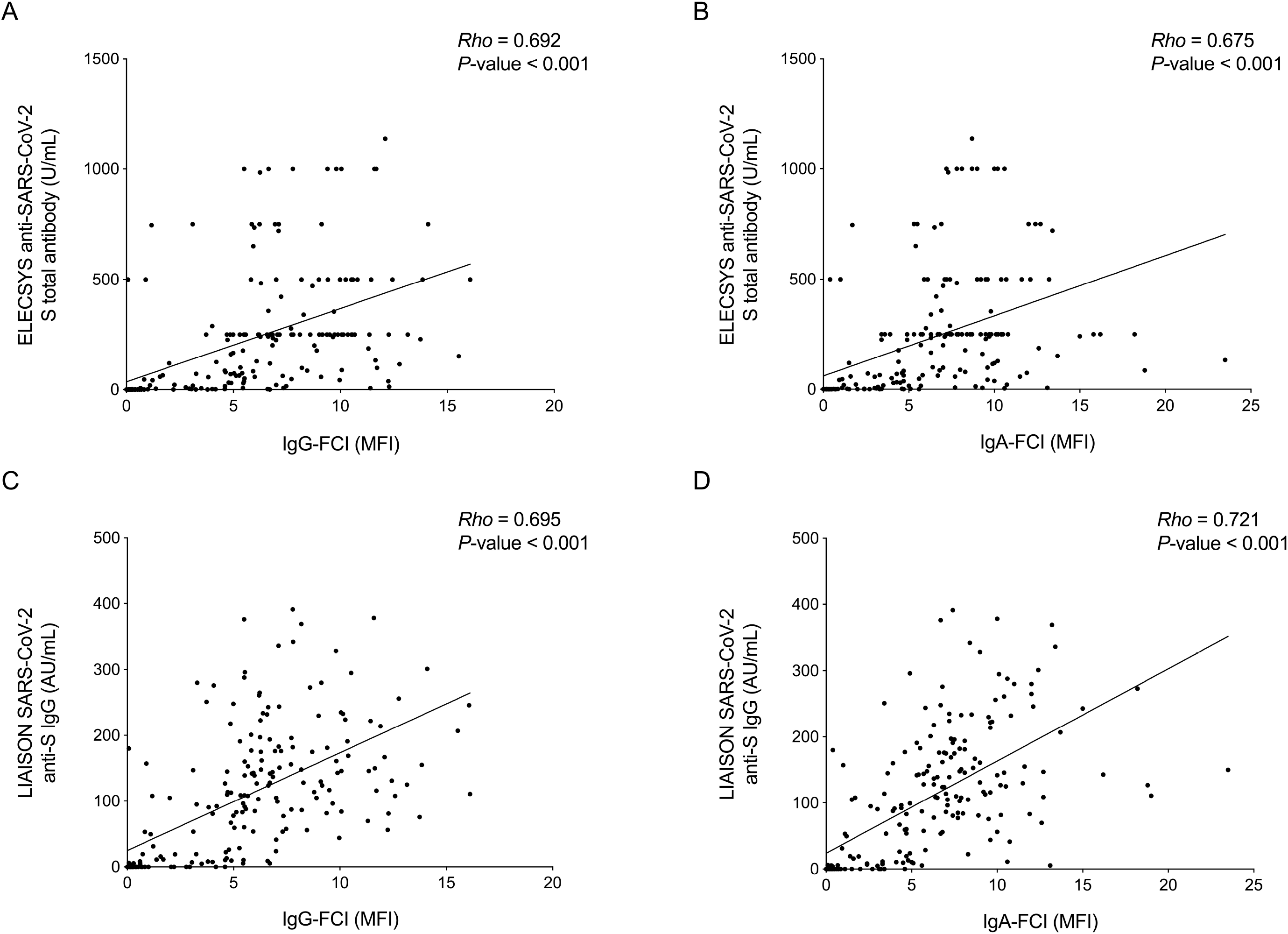
Correlation between serum antibody levels measured by IgG (panels A and C) and IgA (panels B and D) flow cytometry-based immunoassay (FCI) and Elecsys SARS-CoV-2 total antibody electrochemiluminescent assay (Roche diagnostics) and LIAISON® SARS-CoV-2 TrimericS IgG chemiluminescent assay (Diasorin S.p.a). MFI indicates median fluorescence intensity.

### Kinetics of antibody levels measured by different immunoassays

A total of 22 patients belonging to groups I and II had three or more sequential sera available for kinetic analyses. Trajectories of antibody levels delineated by the different immunoassays were categorized as either ascending, descending, ascending to a peak then descending, or fluctuating. As displayed in Table 4, congruent trajectories were seen more frequently for IgG-FCI and IgA-FCI, followed by IgG-FCI or IgA-FCI and IgG measured by Diasorin CLIA. Concordant trajectories were observed commonly for antibodies quantified by Roche ECLIA and Diasorin CLIA; surprisingly, congruent trajectories were seldom seen for IgG or IgA-FCI and total antibodies measured by Roche ECLIA. Fig. 2 exemplifies concordant (2A-D) or divergent (2E-H) antibody trajectories as delineated by the different immunoassays.

**Figure 2.**
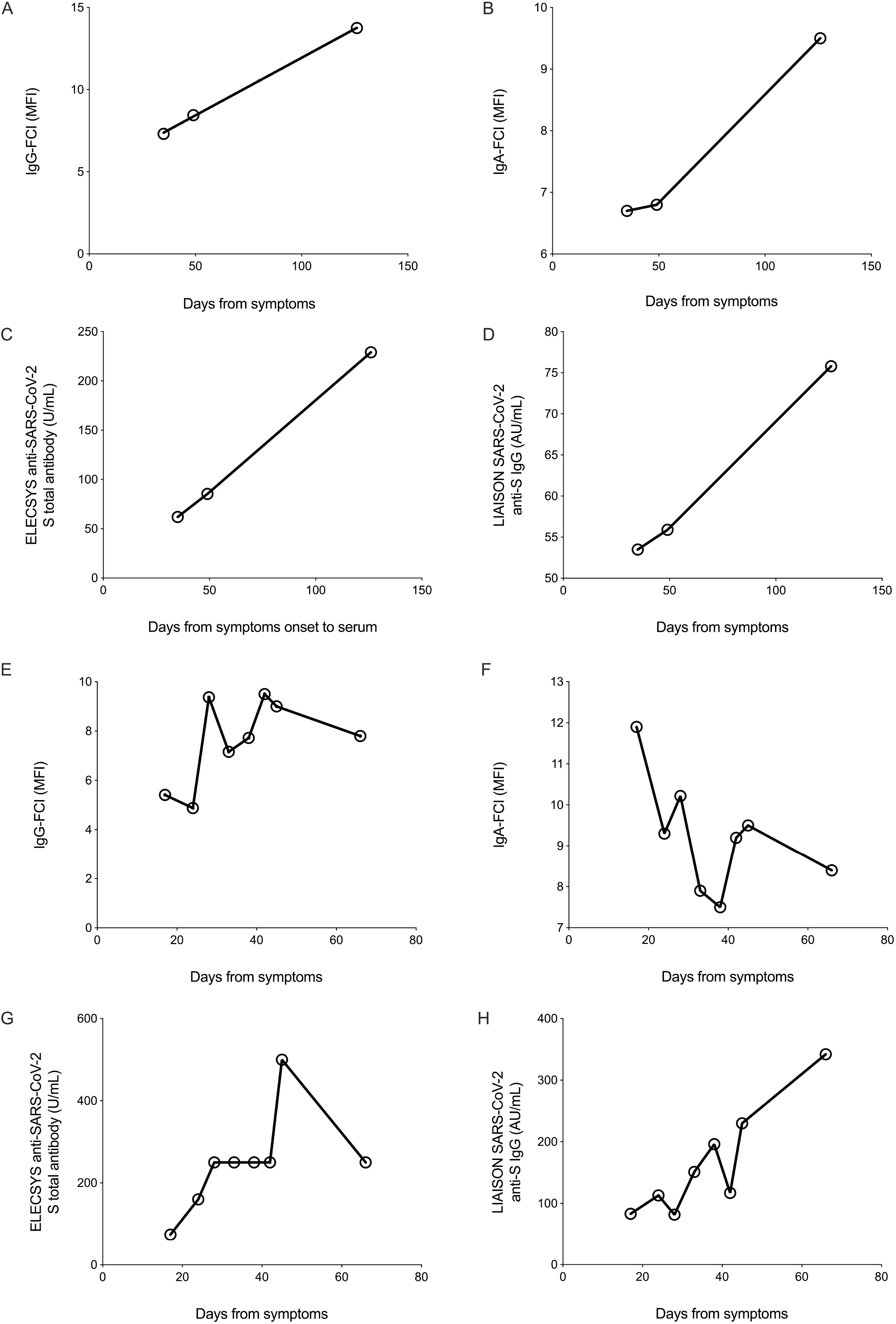
Trajectory of serum/plasma antibody levels as delineated by the immunoassays evaluated in the study, namely IgG or IgA flow cytometry-based immunoassay (FCI), Elecsys SARS-CoV-2 total antibody electrochemiluminescent assay (Roche diagnostics) and LIAISON® SARS-CoV-2 TrimericS IgG chemiluminescent assay (Diasorin S.p.a). A representative example of concordant (A-D) and discordant (E-H) antibody trajectories are shown. MFI indicates median fluorescence intensity.

## DISCUSSION

In this study we compared the performance of an in-house-developed quantitative FCI (12) with the SARS-CoV-2 trimericS-IgG CLIA from Diasorin and Roche RBD-specific IgG/IgM antibody ECLIA for serological diagnosis of SARS-CoV-2 infection in patients with either acute or convalescent COVID-19. All three assays have been reported to measure serum/plasma antibody levels that correlate with those quantified by virus neutralization assays, using either wild type SARS-CoV-2 or lentiviral-S-pseudotyped virions (8-12). Of note, only Roche ECLIA is calibrated to the first WHO International Standard and Reference Panel for anti-SARS-CoV-2 antibody (15). By using a large number of pre-pandemic sera we found the specificity of FCI to be 100%, in agreement with a previous estimate (12). Nevertheless, it must be stressed that we are not certain that sera from individuals with past seasonal coronavirus infection were represented in the panel. The pre-pandemic sera was not run with Roche ECLIA and Diasorin CLIA, and 100% specificity was assumed for both Roche ECLIA as stated by the manufacturer, and Diasorin CLIA as recently reported (9). The main findings of the current study can be summarized as follows. First, direct comparison between IgG-FCI and Diasorin CLIA is biologically straightforward since both assays employ native SARS-CoV-2 S protein as the binding antigen and target the same antibody isotype. Notwithstanding this, both PPA and NPA were below 92% and inter-rater agreement between immunoassays was only moderate (k=0.69). Moreover, correlation between SARS-CoV-2 S IgG levels quantified by both immunoassays was strong (Rho=0.69), but certainly not as strong as anticipated. The lack of full concordance between the results provided by the two assays may relate to subtle differences in the conformation of the binding S protein: whereas in Diasorin CLIA the S protein bound to solid phase exhibits a stable native trimeric conformation, both trimeric and monomeric versions of the S protein were found to be displayed on the surface of transfected Jurkat T cells (12). Furthermore, since SARS-CoV-2-S IgA responses can be documented in the absence of detectable SARS-CoV-2-S IgGs (17), it was not unexpected to observe that PPA decreased whereas NPA increased when IgA FCI results were considered for the analyses, either individually or in combination with IgG ones. Second, despite the fact that Roche ECLIA measures total antibodies (IgG and IgM) binding to the RBD domain of S1 subunit protein instead of the native full-length S protein, we found excellent PPA between the results returned by this assay and by FCI (IgG, IgA or IgG/IgA), ranging between 96.1% to 97.7%, and strong inter-rater agreement (*k* value >0.8), reinforcing the idea that humoral immune response against SARS-CoV-2 following natural infection is mainly directed towards RBD (3-5). In turn, the lower NPA between FCI and Roche ECLIA than between FCI and Diasorin CLIA can be explained by the fact that highly immunogenic B-specific epitopes lie outside the RBD (3). Along with isotype-related differences in the humoral response against SARS-CoV-2, this may also partly account for the suboptimal correlation (Rho, 0.69 for IgG-FCI and Rho, 0.67 for IgA-FCI) between antibody levels quantified by the two immunoassays. Third, both IgA and IgG/IgA-FCI returned more positive results overall than Roche ECLIA and Diasorin CLIA. Interestingly, all three assays, most notably FCI, returned a number of positive results in sera that had scored negative by CLIA assays targeting recombinant S1/S2 subunit proteins or RBD, which were in use for routine diagnosis of SARS-CoV-2 infection at the time of testing request (18). Assuming a specificity of 100% for all assays, these data suggest that the immunoassays evaluated herein, most strikingly FCI, may increase clinical sensitivity of previously marketed assays such as LIAISON^®^ SARS-CoV-2 S1/S2 IgG CLIA and MAGLUMI 2019-nCoV IgG. Fourth, analyzing the kinetics of serum antibody levels in a number of patients with sequential specimens brought the conclusion that the antibody trajectories quantified by the different immunoassays were seldom congruent, with the exception of IgG and IgA determined by FCI assay and to a lesser extent total antibodies quantified by Roche ECLIA and IgG measured by Diasorin CLIA. This observation raises doubts as to the comparability of studies addressing the impact on clinical outcomes of antibody kinetics (i.e. peak levels or the area under a curve), if they employ different immunoassays.

In our view, the main limitations of the current study are the relative small number of specimens included in the evaluation panel and that discrepancies across results returned by the evaluated immunoassays were not resolved by performing antibody neutralization assays, the gold standard for serological diagnosis of SARS-CoV-2 infection (6).

In summary, herein we have shown that a FCI using Jurkat T cells expressing the SARS-CoV-2 native S protein for detection of IgG and IgA-specific antibodies is highly specific and seemingly provides increased clinical sensitivity for diagnosis of SARS-CoV-2 infection when compared to two new-generation immunoassays targeting either the S protein in its trimeric conformation (Diasorin CLIA) or RBD (Roche ECLIA). The assay is easy to perform and standardize; the need for a flow cytometer should not be viewed as a disadvantage compared to high-throughput CLIA assays, as this platform is widely available at immunology and hematology departments in hospitals of all sizes. Further studies evaluating the performance of FCI for documenting seroconversion in vaccinated people are underway.

## Data Availability

The data that support the findings of this study are available on request from the corresponding author.

## ACKNOWLEDGEMENTS

We thank Vitro S.A (Seville, Spain) for providing reagents, flow cytometer equipment and software for the FCI. El Centro Superior de Investigaciones Científicas (CSIC), Spain holds a patent for the method in which the FCI used herein was based upon. FCI [12]. Eliseo Albert holds a Juan Rodés Contract (JR20/00011) from Instituto de Salud Carlos III (Madrid, Spain). Ignacio Torres holds a Río Hortega Contract (CM20/00090) from Instituto de Salud Carlos III (Madrid, Spain).

