## Supplementary material for "Performance Comparison of a Flow Cytometry-based and Two Commercial Chemiluminescent Immunoassays for Detection and Quantification of Antibodies Binding to SARS-CoV-2 Spike Protein": Table 1

| **TABLE 1. Performance comparison between a flow cytometry-based S immunoassay and two commercially-available SARS-CoV-2 chemiluminescent immunoassays** | | |
| --- | --- | --- |
| Assays under comparison (no. of specimens) | Flow cytometry SARS-CoV-2 S assay result (IgG/IgA/IgG+IgA) | |
|  | Positive | Negative |
| Positive by Roche Elecsys® Anti-SARS-CoV-2 S (175) | 168/170/172 | 7/5/3 |
| Negative by Roche Elecsys® Anti-SARS-CoV-2 S (42) | 4/7/7 | 38/35/35 |
| Positive by Liaison® SARS-CoV-2 TrimericS IgG assay (154) | 150/152/153 | 4/2/1 |
| Negative by Liaison® SARS-CoV-2 TrimericS IgG assay (61) | 20/23/24 | 41/38/37 |
| RBD, receptor binding domain; S, spike protein. | | |
