## Supplementary material for "Performance Comparison of a Flow Cytometry-based and Two Commercial Chemiluminescent Immunoassays for Detection and Quantification of Antibodies Binding to SARS-CoV-2 Spike Protein": Table 2

| **TABLE 2. Qualitative results returned by Flow cytometry-based and CLIA assay in specimens from RT-PCR-confirmed SARS-CoV-2 infection testing negative by immunoassays in use at the time of specimen collection^a^** | | | |
| --- | --- | --- | --- |
| Result by corresponding assay | Flow cytometry SARS-CoV-2 S assay (IgG/IgA/IgG+IgA) | Roche Elecsys® Anti-SARS-CoV-2 S^b^ | Liaison® SARS-CoV-2 TrimericS IgG assay^b^ |
| Positive | 18/22/22 | 13 | 5 |
| Negative | 32/28/28 | 34 | 43 |
| ^a^Either lateral flow immunochromatography assay (INNOVITA 2019‐nCoV Ab Test; Beijing Innovita Biological Technology, China), LIAISON SARS-CoV-2 S1/S2 IgG chemiluminescent assay (DiaSorin S.p.A., Saluggia, Italy) or MAGLUMI 2019-nCoV IgG assay (SNIBE – Shenzhen New Industries Biomedical Engineering Co., Ltd, Shenzhen, China).  ^b^2 and 3 specimens could not be analyzed by Liaison and Roche assay, respectively, due to insufficient sample volume. | | | |
