## Supplementary material for "Performance Comparison of a Flow Cytometry-based and Two Commercial Chemiluminescent Immunoassays for Detection and Quantification of Antibodies Binding to SARS-CoV-2 Spike Protein": Table 3

| **TABLE 3. Agreement between qualitative results provided by flow cytometry-based S immunoassay and two commercially-available SARS-CoV-2 CLIA targeting either the trimeric S protein or the receptor binding domain (RBD)** | | | |
| --- | --- | --- | --- |
| Assays under comparison | Percent agreement between qualitative results (95% CI) | | Kappa index (*P* value) |
|  | Positive | Negative |  |
| IgG FCI/Roche Elecsys® Anti-SARS-CoV-2 S | 97.7 (95.4-99.9) | 84.4 (73.8-95.0) | 0.84 (<0.001) |
| IgA FCI/Roche Elecsys® Anti-SARS-CoV-2 S | 96.0 (93.1-98.9) | 87.5 (77.2-97.7) | 0.82 (<0.001) |
| IgG+IgA FCI/Roche Elecsys® Anti-SARS-CoV-2 S | 96.1 (92.2-98.9) | 92.1 (83.5-100) | 0.85 (<0.001) |
| IgG FCI/ Liaison® SARS-CoV-2 TrimericS IgG assay | 88.2 (83.3-93.1) | 91.1 (82.8-99.4) | 0.69 (<0.001) |
| IgA FCI/ Liaison® SARS-CoV-2 TrimericS IgG assay | 86.8 (81.8-91.8) | 95.0 (88.25-100) | 0.68 (<0.001) |
| IgG+IgA FCI/ Liaison® SARS-CoV-2 TrimericS IgG assay | 86.4 (81.4-91.5) | 97.4 (92.3-100) | 0.68 (<0.001) |
| Roche Elecsys® Anti-SARS-CoV-2 S /Liaison® SARS-CoV-2 TrimericS IgG assay | 89.8 (85.3-94.4) | 97.6 (93.0-100) | 0.76 (<0.001) |
| FCI, flow cytometry immunoassay. | | |  |
