## Supplementary material for "Performance Comparison of a Flow Cytometry-based and Two Commercial Chemiluminescent Immunoassays for Detection and Quantification of Antibodies Binding to SARS-CoV-2 Spike Protein": Table 4

| **TABLE 4. Comparison of trajectories of serum antibody levels as delineated by immunoassays under comparison** | |
| --- | --- |
| **Concordant antibody trajectories as delineated by SARS-CoV-2 immunoassays** | **Nº of patients** |
| All | 1 |
| IgG-FCI/IgA-FCI | 11 |
| IgG-FCI/Roche ECLIA | 0 |
| IgA-FCI/Roche ECLIA | 0 |
| IgG-FCI/IgA-FCI/Roche ECLIA | 0 |
| IgG-FCI/Diasorin CLIA | 2 |
| IgA-FCI/Diasorin CLIA | 2 |
| IgG-FCI/IgA-FCI/Diasorin CLIA | 3 |
| Roche ECLIA/Diasorin CLIA | 8 |
| IgG-FCI/Diasorin CLIA/Roche ECLIA | 3 |
| IgA-FCI/Diasorin CLIA/Roche ECLIA | 0 |
| IgG-FCI/IgA-FCI/Diasorin CLIA/Roche ECLIA | 1 |
| None | 2 |
| CLIA, chemiluminescent immunoassay (Liaison® SARS-CoV-2 TrimericS IgG assay); ECLIA, electrochemiluminescent immunoassay (Roche Elecsys® Anti-SARS-CoV-2 S assay); FCI, flow cytometry-based immunoassay; | |
